# Community adherence to the second dose of measle and rubella: a cross-sectional survey in a rural health district of Cameroon

**DOI:** 10.1101/2025.06.07.25329191

**Authors:** Guy Stéphane Nloga, Adidja Amani, Florence Kissougle Nkongo, Roger Mathurin Mfoula Angoula, Fabrice Zobel Lekeumo Cheuyem

## Abstract

**Background:** Measles remains a significant public health challenge in sub-Saharan Africa, including Cameroon, where vaccination coverage falls below the WHO target of 95%. The second dose of the measles-rubella vaccine (MCV2) is critical for strengthening immunity, yet adherence remains low in rural areas. This study aimed to assess MCV2 coverage and identify factors influencing vaccination uptake in the Ngog-Mapubi Health District of Cameroon.

**Methods:** A community-based cross-sectional survey was conducted in March 2024, involving 140 parents or guardians of children aged 15–23 months. Data were collected using a pretested questionnaire, covering sociodemographic characteristics, vaccination status, and barriers to uptake. Logistic regression was employed to identify determinants of MCV2 vaccination. Data were analyzed using R statistics version 4.4.2. A *p*-value 11 0.05 was considered statistically significant.

**Results:** The study revealed an MCV2 vaccination coverage of 46.4% (95% CI: 38.0–55.0). Among unvaccinated children, 85.3% (95% CI: 75.3–92.4) of caregivers expressed willingness to vaccinate, indicating high acceptance but persistent barriers. Distance to health facilities (43%), lack of information about vaccine availability (37%), and missed opportunities during health visits (25%) were the most cited obstacles. Awareness of MCV2 was high (89%), but knowledge gaps persisted, with 61% of respondents demonstrating poor understanding of vaccination schedules. Multivariate analysis identified the absence of advanced vaccination strategies (e.g., outreach programs) as the strongest predictor of non-vaccination (aOR = 7.15; 95% CI: 3.19–17.2; *p* < 0.001). Sociodemographic factors like single/widowed marital status (cOR = 3.15; *p* = 0.025) and student occupation (cOR = 10.0; *p* = 0.048) were also associated with lower uptake at univariate analysis. Geographic disparities were notable, with coverage below 10% in three health areas (Makak, Mbebe-kikot, Ntouleng). Healthcare workers were the primary information source (93.6%).

**Conclusion:** Despite the high MCV2 acceptance rate, low MCV2 coverage in this rural district underscores the need for improved access through advanced vaccination strategies and targeted community education. Addressing structural and informational barriers is essential to achieving equitable immunization coverage.

## Introduction

In 2015, the United Nations General Assembly established the Sustainable Development Goals, which mandate that by 2030, all countries reduce their under-5 mortality rate to no more than 25 deaths per 1,000 live births and their neonatal mortality rate to no more than 12 deaths per 1,000 live births [1]. To achieve this global health objective, vaccines are one of the most powerful tools used to prevent disease among children and save millions of lives each year [2]. Since the launch of global vaccination programs in 1974, an estimated 154 million deaths have been prevented (146 millions of these among children under 5 years), including 101 million infants under 1 year [3]. There are vaccines for over 30 life-threatening diseases, enabling people of all ages to live longer and healthier by protecting them against illnesses such as diphtheria, tetanus, pertussis (whooping cough), pneumonia, diarrhea, and measles [2].

Measles remains a highly contagious viral disease that primarily affects children, characterized by severe symptoms including high fever, anorexia, cough, rash, and potentially fatal complications such as pneumonia and blindness [4]. In the pre-vaccine era prior to 1963, global measles epidemics occurred every few years causing an estimated 2.6 million deaths annually worldwide [5]. Sub-Saharan Africa continues to experience the world’s highest measles burden, with 4.23 deaths and 355.68 disability-adjusted life years lost per 100,000 population [6]. The region witnessed a dramatic resurgence in 2022, particularly in Ethiopia, Somalia, and the Democratic Republic of Congo [7]. The COVID-19 pandemic severely disrupted measles prevention efforts by overwhelming healthcare systems and suspending routine immunization programs [8,9]. As a result, 23 million children in the region missed essential vaccines, including the measles containing vaccine (MCV), leaving them vulnerable to preventable outbreaks [5].

Despite global progress in measles control, Cameroon continues to face recurrent outbreaks affecting several regions, including the Centre, Littoral, South-West, and Northern regions [6,10]. As of 26 November 2023, 6,054 measles cases and 31 measles-related deaths had been confirmed since the beginning of the year, with the Centre region being the most affected [11]. To strengthen immunity in children under five, WHO initially recommended in 2009 that the second measles vaccine dose (MCV2) be introduced into routine immunization only after achieving ≥80% MCV1 coverage nationally for three consecutive years [12]. However, this policy was updated in April 2017 to recommend MCV2 inclusion in all national vaccination schedules, regardless of MCV1 coverage levels [13].

Cameroon’s vaccination coverage (77% in 2017) remains below the WHO’s 95% target, with high regional disparities [6]. Estimates have decreased further to 27.9%, according to recent 2022 reports from the Centre region [14]. These coverage gaps create outbreak-prone areas, particularly in rural areas, which typically experience disparities in health workforce availability compared to urban areas [15]. This coverage depends on various factors including the level of acceptance or hesitancy towards vaccines in the community. To this regard, study reports identified several factors that could influence MCV uptake. Such factors include education which plays a crucial role in vaccination uptake, with lower maternal or caregiver educational attainment consistently reducing the likelihood of a child being vaccinated, while higher education levels progressively increase vaccine acceptance [16]. Significant barriers to healthcare access also impede immunization efforts; longer travel times to health facilities and prior experiences of vaccine stockouts actively deter future visits [16,17]. Furthermore, home deliveries, as opposed to births in health facilities, are associated with reduced subsequent immunization rates [16]. Active engagement with healthcare services positively influences vaccination compliance; mothers attending postnatal care visits show higher adherence. Correct knowledge of required vaccination visits is also vital for improving adherence [16,18]. Within household dynamics, families with multiple young children are less likely to be vaccinated [17]. Critically, knowledge gaps contribute to lower uptake, as a limited understanding of vaccine-preventable diseases and unawareness of required measles vaccine doses decrease compliance [18]. Structural challenges, such as rural residence, correlate with reduced access to vaccination services, and younger caregivers tend to have lower vaccination rates than their older counterparts. Finally, economic status are significant factors, with lower household income decreasing vaccination likelihood, and competing priorities in large families potentially diverting resources from preventive care [17]. There is limited evidence on MCV2 vaccination uptake and related factors in Cameroon [6,14]. The present study aimed to provide an insight of MCV2 coverage and factors influencing its consistent uptake in a rural Health District (HD) of the Centre Region of Cameroon.

## Methods

### Study design and period

We conducted in March 2024 a community based cross-sectional study with descriptive and analytical purpose in health areas of the Ngog-Mapubi HD in the Centre region of Cameroon.

### Study setting

The HD represents the operational level of the health system pyramid in Cameroon and constitutes the unit of implementation of national priority health programs. The Ngog-Mapubi is located in the Nyong-et-Kéllé Division, and it is one of the 32 HDs in the Centre Region. The district comprises five subdivisions including Bot-Makak, Dibang, Matomb, Ngog-Mapubi, and Nguibassal. Its population in 2024 is estimated at 59,545, distributed across 14 health areas. The HD’s healthcare map encompasses 28 health facilities including one District Hospital, four Subdivisional Medical Centers, 15 Integrated Health Centers, three private secular health facilities, and five private religious health facilities. It is a rural HD with a predominantly Basa’a ethnic group whose main activity is agriculture.

### Study participants

This study targeted parents and legal guardian of children aged 15-23 months in the community of the Ngog-Mapubi HD. All eligible community members who accepted to participate in the study and provided an inform consent were included in the study.

### Sampling

All 14 health areas of the HD were sampled for the study. The minimum sample size, calculated using the Cochrane formula, was 303 households, based on an estimated MCV2 coverage proportion of 27,9% [14]. To distribute the number of households across the different health areas, we proportionately allocated the sample size among them using the following formula: 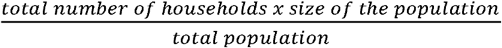. Systematic random sampling was used to select the households to be surveyed. The sampling interval (increment) observed was 2. The initial direction was determined by throwing a pen in the air from a central point in the neighborhood, typically represented by the chief’s house in each surveyed village. In each selected household, one eligible child was randomly chosen from all children aged 15-23 months within that household for the assessment of vaccination status and the collection of other relevant information.

### Data collection tool and procedures

A pretested data collection tool was developed for the purposes of this study. It collected data related to sociodemographic characteristics, MCV2 vaccination coverage and acceptance, and common barriers to vaccination. Data collection was carried out by trained community health workers from the Ngog-Mapubi HD.

### Variable and operational definition

A child was considered vaccinated based on the parent or legal guardian’s oral report or the presentation of the vaccination booklet. Vaccine acceptance was defined as answering “yes” to the question, “Are you willing to get your eligible child vaccinated with the MCV2 if it was proposed to you?”

The dependent variable for the logistic regression was the child’s MCV2 vaccination status and the risk was having in the household an eligible unvaccinated child against the disease. Independent variables included the parent or legal guardian’s marital status, education level, ethnic group, religion, the number of children in the household, and monthly average income. We also assessed the participants’ awareness of the MCV2 vaccine and their knowledge regarding this specific vaccine.

### Data processing and analysis

Data were recorded, processed and then analyzed using R statistics version 4.4.2 [19]. The Fisher exact test was used to compare proportions. Non-normal continuous variables were described using median and interquartile ranges. A score of 1 was attributed to each good answer on the three questions assessing knowledge. The participant knowledge was categorized as poor (0 to 1/3), faire (2/3), and good (3/3). Simple and multiple binomial logistic regressions were used to identify determinants of vaccine uptake, and adjusted Odds Ratios were used to establish the strength of the association between the variables and to eliminate potential confounding factors. The selection of predictors that best fit the model was done stepwise using the Akaike Information Criterion (AIC). The model with the lowest index was then selected [20]. A *p*-value (*p*) < 0.05 was considered statistically significant, and confidence intervals (CI) were estimated at the 95% confidence level.

## Results

A total of 390 eligible households were reached during this study. A total of 167 parents or legal guardians were available for the interview among whom 140 accepted to participate (response rate 84%).

### Sociodemographic characteristics of respondents

All fourteen health areas belonging to the HD were sampled. Most of the respondents were aged 20-39 years (82%), had a secondary study level (62%), and the female gender was the most represented (85%). They were mostly from the Centre Region (94%), belonging to the local tribe (Basa’a = 84%), Catholic (63%), housekeepers by occupation (55%), and with less than 50,000 XFA as average monthly income (86%) (Table 1).

**Table 1.**
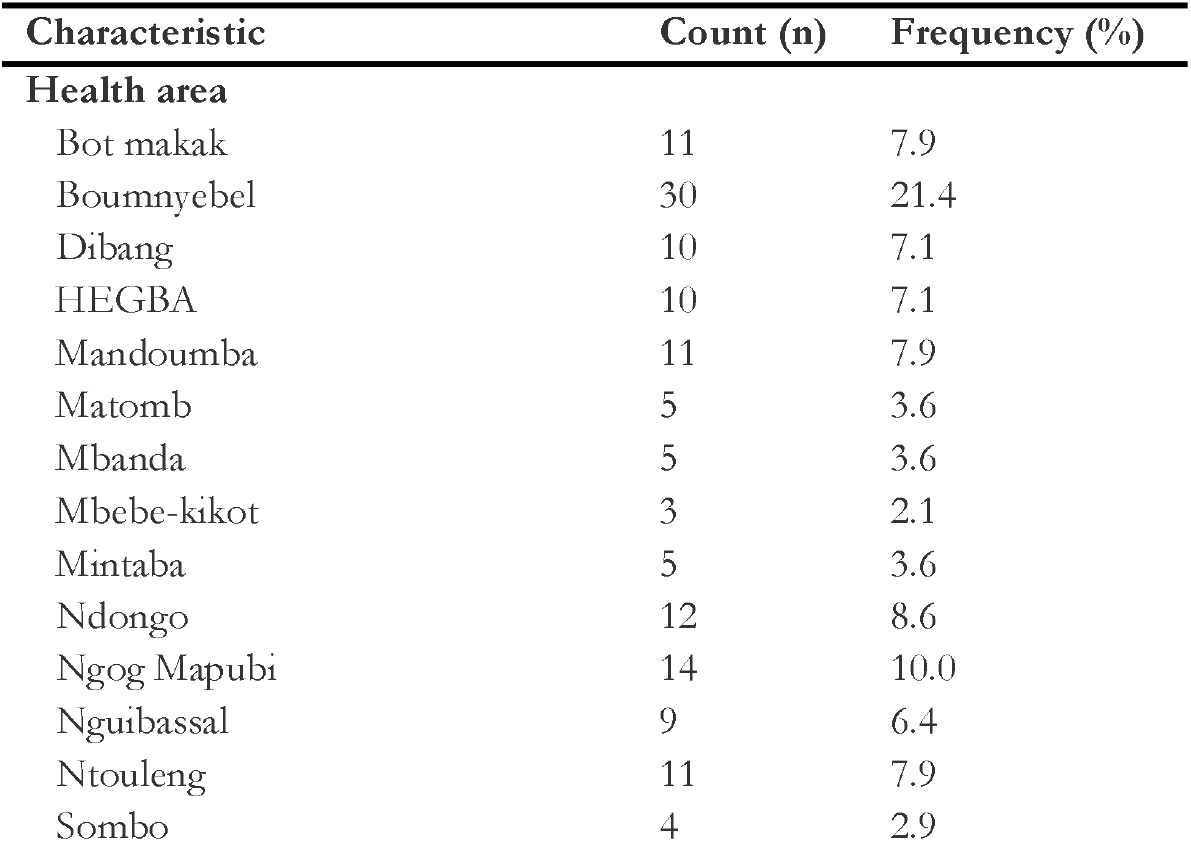
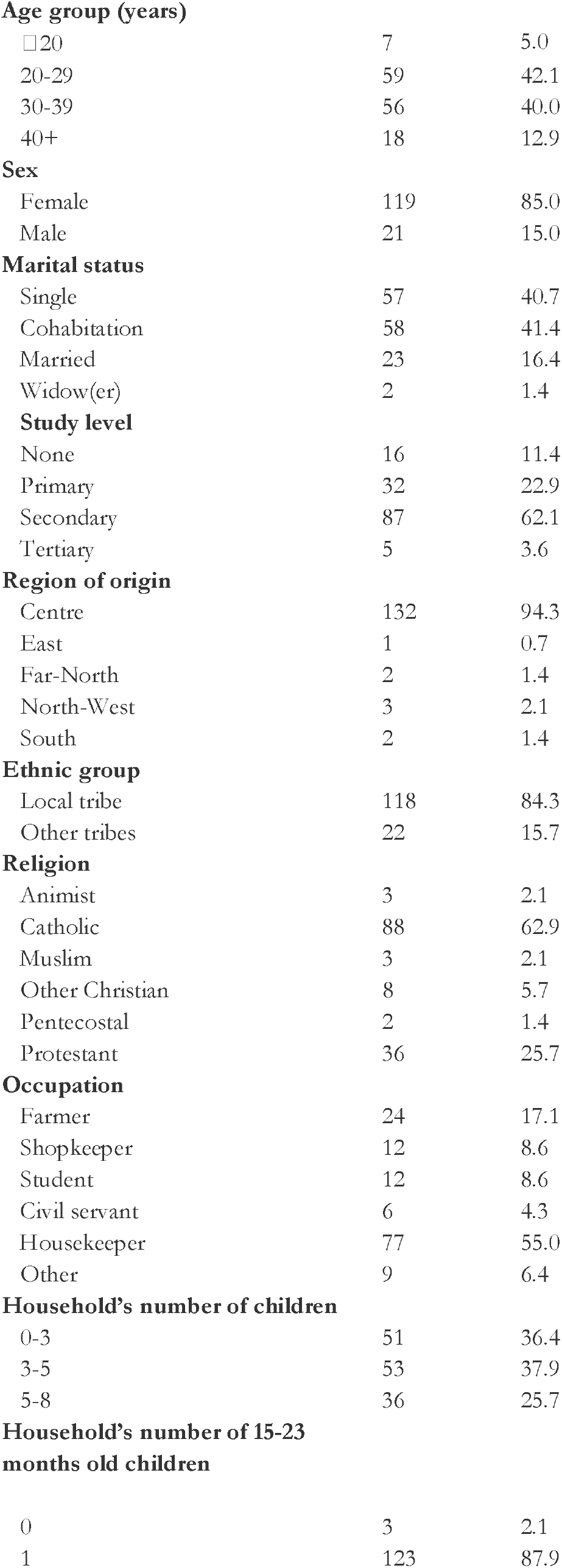
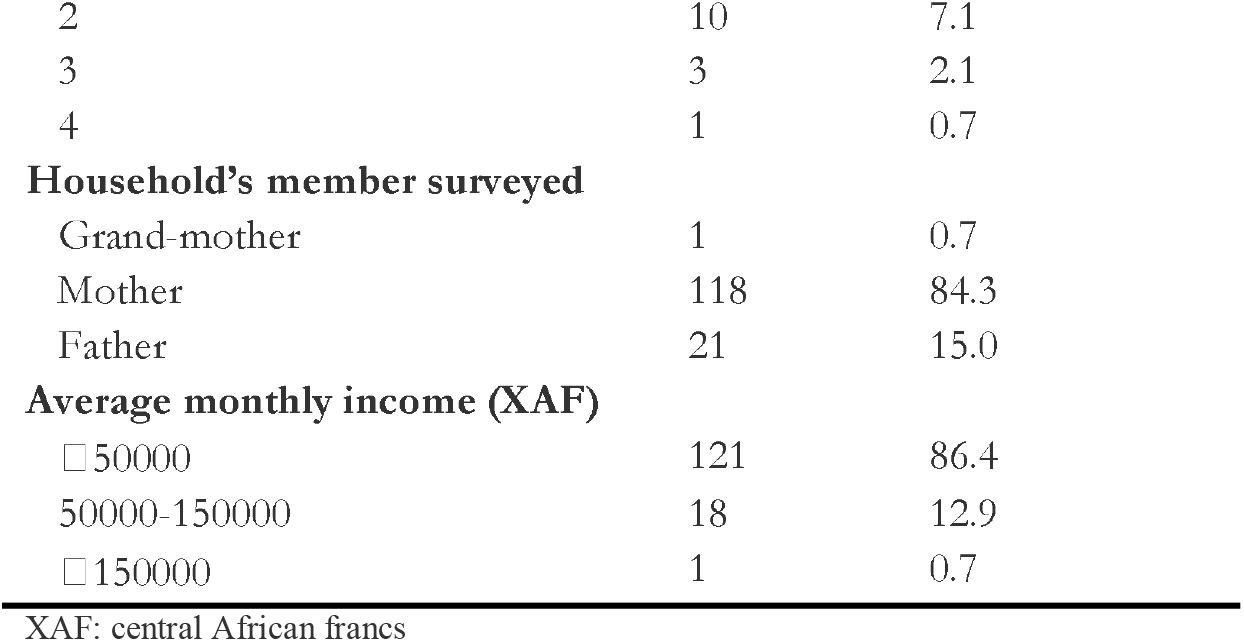
Sociodemographic profile of study participants, Ngog-Mapubi HD, Cameroon (n = 140)

| Characteristic | Count (n) | Frequency (%) |
| --- | --- | --- |
| **Health area** |  |  |
| Bot makak | 11 | 7.9 |
| Boumnyebel | 30 | 21.4 |
| Dibang | 10 | 7.1 |
| HEGBA | 10 | 7.1 |
| Mandoumba | 11 | 7.9 |
| Matomb | 5 | 3.6 |
| Mbanda | 5 | 3.6 |
| Mbebe-kikot | 3 | 2.1 |
| Mintaba | 5 | 3.6 |
| Ndongo | 12 | 8.6 |
| Ngog Mapubi | 14 | 10.0 |
| Nguibassal | 9 | 6.4 |
| Ntouleng | 11 | 7.9 |
| Sombo | 4 | 2.9 |
| **Age group (years)** |  |  |
| □20 | 7 | 5.0 |
| 20-29 | 59 | 42.1 |
| 30-39 | 56 | 40.0 |
| 40+ | 18 | 12.9 |
| **Sex** |  |  |
| Female | 119 | 85.0 |
| Male | 21 | 15.0 |
| **Marital status** |  |  |
| Single | 57 | 40.7 |
| Cohabitation | 58 | 41.4 |
| Married | 23 | 16.4 |
| Widow(er) | 2 | 1.4 |
| **Study level** |  |  |
| None | 16 | 11.4 |
| Primary | 32 | 22.9 |
| Secondary | 87 | 62.1 |
| Tertiary | 5 | 3.6 |
| **Region of origin** |  |  |
| Centre | 132 | 94.3 |
| East | 1 | 0.7 |
| Far-North | 2 | 1.4 |
| North-West | 3 | 2.1 |
| South | 2 | 1.4 |
| **Ethnic group** |  |  |
| Local tribe | 118 | 84.3 |
| Other tribes | 22 | 15.7 |
| **Religion** |  |  |
| Animist | 3 | 2.1 |
| Catholic | 88 | 62.9 |
| Muslim | 3 | 2.1 |
| Other Christian | 8 | 5.7 |
| Pentecostal | 2 | 1.4 |
| Protestant | 36 | 25.7 |
| **Occupation** |  |  |
| Farmer | 24 | 17.1 |
| Shopkeeper | 12 | 8.6 |
| Student | 12 | 8.6 |
| Civil servant | 6 | 4.3 |
| Housekeeper | 77 | 55.0 |
| Other | 9 | 6.4 |
| **Household's number of children** |  |  |
| 0-3 | 51 | 36.4 |
| 3-5 | 53 | 37.9 |
| 5-8 | 36 | 25.7 |
| **Household's number of 15-23 months old children** |  |  |
| 0 | 3 | 2.1 |
| 1 | 123 | 87.9 |
| 2 | 10 | 7.1 |
| 3 | 3 | 2.1 |
| 4 | 1 | 0.7 |
| **Household's member surveyed** |  |  |
| Grand-mother | 1 | 0.7 |
| Mother | 118 | 84.3 |
| Father | 21 | 15.0 |
| **Average monthly income (XAF)** |  |  |
| □ 50000 | 121 | 86.4 |
| 50000-150000 | 18 | 12.9 |
| □ 150000 | 1 | 0.7 |

XAF: central African francs

The median distance between the household and the health facility providing immunization service was 4 km (Q1-Q2: 2-8km) with a minimum of 111 km and a maximum of 50 km.

### MCV2 vaccination coverage and acceptance

Nearly half of eligible children had received the second dose of the MCV (46.4%; 95% CI: 38.0-55.0), and most of the respondents whose children were not yet vaccinated declared accepting to vaccinate their children (85.3%; 95% CI: 75.3-92.4). Most of the respondents reported their adherence to expanded program of immunization implemented (EPI), by usually getting their kids vaccinated in the nearby immunization service (93.6%; 95% CI: 88.1-97.0). The lowest vaccine coverage was significantly observed in both Makak, Mbebe-kikot, and Ntouleng (<10%) health areas, while the lowest acceptance rate among households with unvaccinated eligible children was significantly recorded in the Nguibassal (0%) health area (Supplementary Tables 1, 2 and 3).

**Table 2.** Knowledge related to MCV2 among community members in Ngog-Mapubi HD, Cameroon (n = 140)

| Knowledge | Count (n) | Frequency (%) |
| --- | --- | --- |
| **Period of the EPI eligibility (months)** |  |  |
| 0-3 | 7 | 5.0 |
| 0-6 | 50 | 35.7 |
| 0-23 (*Yes*) | 10 | 7.1 |
| 0-59 | 73 | 52.1 |
| **Required dose of MCV** |  |  |
| 1 | 11 | 7.9 |
| 2 (*Yes*) | 58 | 41.4 |
| 3 | 5 | 3.6 |
| 4 | 2 | 1.4 |
| I don't know | 64 | 45.8 |
| **Best time to receive the MCV2 (months)** |  |  |
| 3 | 8 | 5.7 |
| 6 | 5 | 3.6 |
| 9 | 27 | 19.3 |
| 12 | 27 | 19.3 |
| 15 (*Yes*) | 73 | 52.1 |
| **Overall level of knowledge<sup>†</sup>** |  |  |
| Poor | 85 | 60.7 |
| Fair | 29 | 20.7 |
| Good | 26 | 18.6 |

EPI: expanded program of immunization in Cameroon; MCV2: second dose of measles and rubella vaccine;
<sup>†</sup>Good: score =3/3=100%; Fair: score=2/3=67%; Poor: score=0-1/3=0-33%

**Table 3.** Univariate and multivariate binary logistic regression of parameters associated with MCV2 non-vaccination in the Ngog-Mapubi HD, Cameroon (n=140)

| Factor | Vaccine uptake<br>n (%) |  | cOR | p-value | aOR | 95% CI limits |  | p-value |
| --- | --- | --- | --- | --- | --- | --- | --- | --- |
|  | Yes<br>65 (46) | No<br>75 (54) |  |  |  | Lower | Upper |  |
| **Marital status** |  |  |  |  |  |  |  |  |
| Married | 15 (65.2) | 8 (34.8) | 1 |  |  |  |  |  |
| Cohabitation | 28 (48.3) | 30 (51.7) | 2.01 | 0.172 |  |  |  |  |
| Single/Widow(er) | 22 (37.3) | 37 (62.7) | 3.15 | 0.025 |  |  |  |  |
| **Study level** |  |  |  |  |  |  |  |  |
| None | 6 (37.5) | 10 (62.5) | 1 |  | 1 |  |  |  |
| Primary | 12 (37.5) | 20 (62.5) | 1.00 | >0.999 | 1.36 | 0.34 | 5.43 | 0.660 |
| Secondary | 46 (52.9) | 41 (47.1) | 0.53 | 0.263 | 0.52 | 0.14 | 1.75 | 0.297 |
| Tertiary | 1 (20.0) | 4 (80.0) | 2.40 | 0.477 | 2.26 | 0.18 | 59.1 | 0.554 |
| **Occupation** |  |  |  |  |  |  |  |  |
| Civil servant | 4 (66.7) | 2 (33.3) | 1 |  |  |  |  |  |
| Farmer | 16 (66.7) | 8 (33.3) | 1.00 | >0.999 |  |  |  |  |
| Shopkeeper | 8 (66.7) | 4 (33.3) | 1.00 | >0.999 |  |  |  |  |
| Housekeeper | 32 (41.6) | 45 (58.4) | 2.81 | 0.249 |  |  |  |  |
| Student | 2 (16.7) | 10 (83.3) | 10.0 | 0.048 |  |  |  |  |
| Other | 3 (33.3) | 6 (66.7) | 4.00 | 0.215 |  |  |  |  |
| **Ethnic group** |  |  |  |  |  |  |  |  |
| Other tribes | 11 (50.0) | 11 (50.0) | 1 |  | 1 |  |  |  |
| Local tribe | 54 (45.8) | 64 (54.2) | 1.19 | 0.715 | 0.89 | 0.31 | 2.48 | 0.820 |
| **Religion group** |  |  |  |  |  |  |  |  |
| Christian | 64 (47.8) | 70 (52.2) | 1 |  |  |  |  |  |
| Muslim | 1 (33.3) | 2 (66.7) | 1.83 | 0.626 |  |  |  |  |
| Animist | 0 (0.0) | 3 (100.0) | NA |  |  |  |  |  |
| **Household's number of children** |  |  |  |  |  |  |  |  |
| 0-3 | 24 (47.1) | 27 (52.9) | 1 |  | 1 |  |  |  |
| 3-5 | 20 (37.7) | 33 (62.3) | 1.47 | 0.337 | 1.31 | 0.52 | 3.33 | 0.561 |
| 5-8 | 21 (58.3) | 15 (41.7) | 0.63 | 0.301 | 0.79 | 0.30 | 2.22 | 0.660 |
| **Household's monthly average income** |  |  |  |  |  |  |  |  |
| □50000 | 54 (44.6) | 67 (55.4) | 1 |  |  |  |  |  |
| 50000-150000 | 10 (55.6) | 8 (44.4) | 0.64 | 0.388 |  |  |  |  |
| □150000 | 1 (100.0) | 0 (0.0) | NA | 0.987 |  |  |  |  |
| **Ever heard of MCV2** |  |  |  |  |  |  |  |  |
| Yes | 65 (52.0) | 60 (48.0) | 1 |  |  |  |  |  |
| No | 0 (0.0) | 15 (100.0) | NA |  |  |  |  |  |
| **Good knowledge** |  |  |  |  |  |  |  |  |
| Yes | 15 (57.7) | 11 (42.3) | 1 |  |  |  |  |  |
| No | 50 (43.9) | 64 (56.1) | 1.75 | 0.205 |  |  |  |  |
| **Advance strategy offered** |  |  |  |  |  |  |  |  |
| Yes | 54 (63.5) | 31 (36.5) | 1 |  |  |  |  |  |
| No | 11 (20.0) | 44 (80.0) | 6.97 | <0.001 | 7.15 | 3.19 | 17.2 | <0.001 |

cOR: crude Odds Ratio; aOR: adjusted Odds Ratio; CI: confidence interval; MCV2: second dose of the measles and rubella

### Output of MCV2 communication activities

Most of the respondents (89%) reported having heard about MCV2, and there was no significant association with sociodemographic characteristics (Supplementary Tables 4 and 5).

Despite this high reported exposure, most of the respondents had poor knowledge (61%) regarding MCV2 (Table 2).

The most commonly reported communication channels included healthcare workers during and outside vaccination sessions (Fig. 1).

**Fig. 1.**
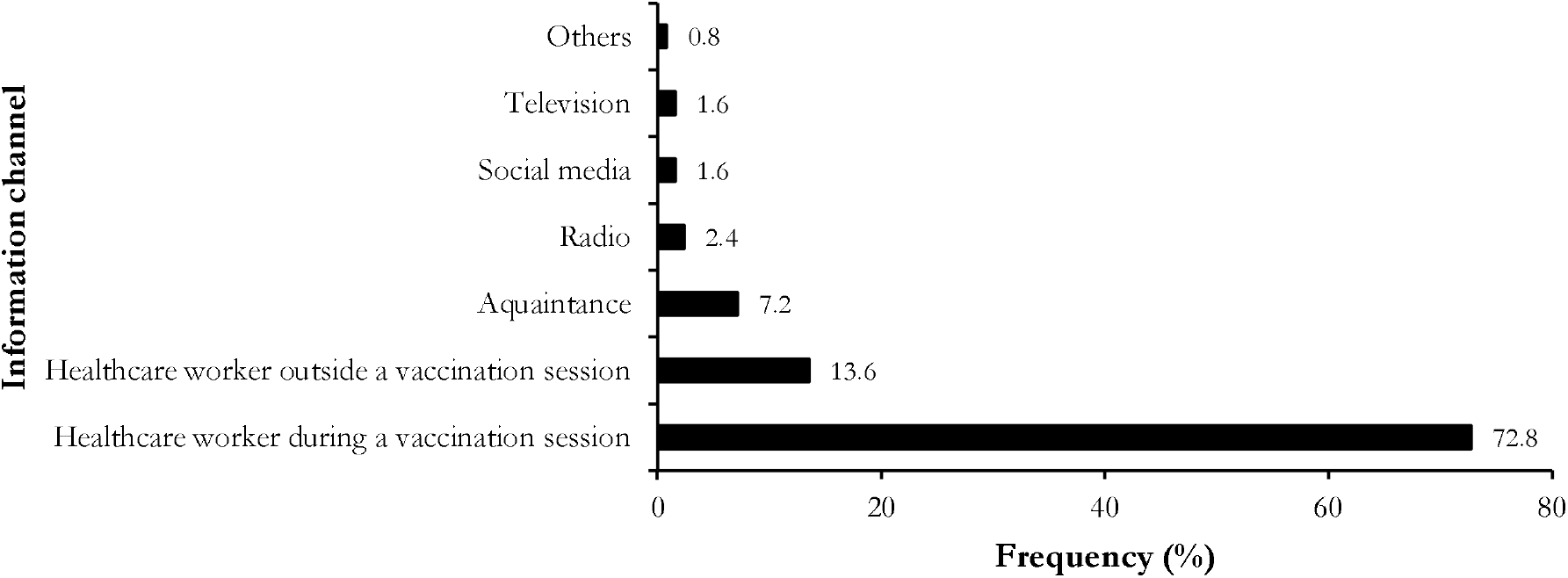
Reported information channel on MCV2 among households’ members in the Ngog-Mapubi HD, Cameroon (n = 125)

The most commonly reported barriers to MCV2 vaccination included the distance to the immunization service (43%), the lack of information about the administration of the vaccination in the health facility (37%) and the fact that some eligible children were brought to the health facility but return back without receiving the vaccine (25%) (Fig. 2). The long distance was also mentioned as the main barrier to compliance with the routine immunization service for children (Supplementary Fig. 1).

**Fig. 2.**
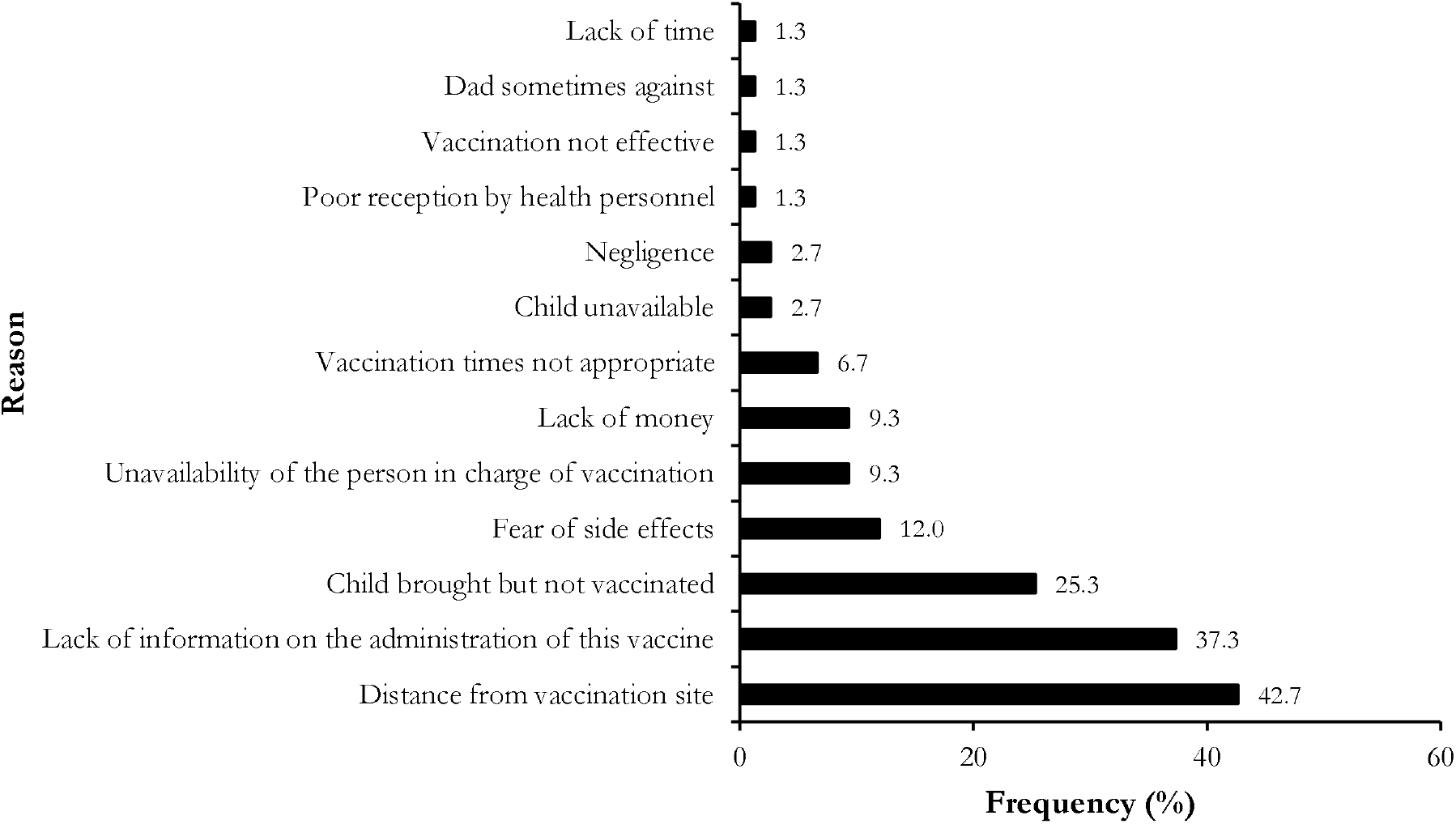
Reported barriers to the vaccination of MCV2 eligible children in Ngog-Mapubi HD, Cameroon (n = 75)

### Factor associated with MCV2 uptake

At univariate analysis the marital status (single/widow(er)), the occupation (student) and the implementation of advance strategy were significantly associated with vaccine uptake. However, at multivariate analysis households whose members reported the absence of advance strategy offer were 7 times more likely to not have their children receive the MCV2 (aOR = 7.15; 95% CI: 3.19-17.2; *p* ⍰ 0.001) (Table 3).

## Discussion

Our study aimed to identify the factors of community non-adherence to the second dose of MCV2 in a rural HD of Cameroon.

The results revealed that 46.4% of eligible children received this antigen, which is far below the expected vaccination coverage of 82% [21]. Multivariate analysis shows that, among the variables studied, the absence of an advanced strategy offer constitutes the main determinant of non-vaccination. However, certain sociodemographic characteristics also showed a significant association or notable trends deserving discussion.

The factor most strongly associated with non-vaccination remains the absence of an advanced strategy offer (aOR = 7.15; 95% CI: 3.19–17.2; *p* < 0.001). This result is similar to those of Nguenga *et al*. [22] and Russo *et al*. [23], who found distances of more than 5 km, as factors of non-adherence to children’s vaccination schedules. This data underscores the operational importance of organizing vaccination services. In rural areas, where geographical accessibility is a major obstacle, advanced strategies are essential to reach the most remote populations. Their absence increases the risk of non-vaccination sevenfold, illustrating the determining role of structural healthcare provision.

The education level of the respondents, although not significantly associated with vaccination in our multivariate model, reveals a trend in which individuals responsible for vaccinating children with a secondary or higher education level showed higher vaccination rates than those with no education or only primary education. This observation corroborates data from the literature, which establish a link between education level and the use of preventive health services, particularly concerning the understanding of awareness messages and adherence to the recommendations of the EPI [16,17].

Regarding marital status, children with single or widowed parents had a significantly higher risk of non-vaccination compared to those living in households with married parents (cOR = 3.15; *p* = 0.025). These findings are corroborated by the study of Yakum *et al*. [24], which revealed that children whose mothers share the same home with their biological father are more likely to be compliant with the EPI schedule. This result suggests that marital stability could promote better planning and improved access to vaccination services, particularly through shared parental responsibilities and increased social support.

Concerning the occupation of the respondents, we noted that student status of the parent is strongly associated with non-vaccination (cOR = 10.0; *p* = 0.048). This result can be explained by the fact that students, generally young, may prioritize infant health obligations less. Conversely, homemakers, although available, did not have significantly better rates, suggesting that other factors such as personal motivation, level of information, or decision-making autonomy might play a role. These results differ from those of Legesse *et al*., whose study showed that parents responsible for vaccinating children whose occupation was farming were 1.7 times more likely to have their children fully vaccinated [25].

Religious affiliation did not emerge as an associated factor in the present study. However, it was associated with incomplete child vaccination in the study by Douba *et al*. [26]. Indeed, in that study, children of mothers of Christian and/or Muslim faith had a reduced risk of being incompletely vaccinated compared to those of animist or non-religious mothers. However, it is worth be noted that in our study, the three respondents who declared themselves animists had not vaccinated their children, although the size of this sample is insufficient to draw statistical conclusions. Future qualitative investigations could explore these cultural aspects further.

Household size, particularly in terms of the number of children, was found to be non-significant. However, households with between five and eight children showed relatively higher vaccination rates, which could reflect greater experience or familiarity with vaccination campaigns.

Regarding monthly income, although no statistically significant association was observed, households with higher incomes showed a trend towards better vaccination coverage. This trend is similar to the results of Legesse *et al*. [25] and Yakum *et al*. [27], who found that households with a high average income were 3 times more likely to have their children fully vaccinated than households with a low average income. This finding is consistent with data indicating that financial constraints can be indirect obstacles to vaccination, particularly due to the inability to travel to health centers.

Awareness of MCV2, measured by having heard about the vaccine, was reported among all respondents who had vaccinated their children. Conversely, none of the 15 respondents who reported never having heard of MCV2 had vaccinated their children, highlighting the crucial importance of information in compliance with vaccination program.

## Limitations

This study has some limitations. Firstly, the cross-sectional design does not allow for establishing a causal relationship between the studied factors and non-vaccination. Secondly, the sample size was not large enough to detect the significance of some variables declared statistically non-significant. Thirdly, the data rely on participants’ declarations, which expose them to recall bias or social desirability bias. Furthermore, the sample, though representative of the Ngog-Mapubi district, might limit the generalization of results to other geographical or sociocultural contexts. Finally, some potentially influential factors, such as the quality of healthcare provision or the attitudes of healthcare professionals, were not explored.

## Conclusions

Despite the high acceptance rate, the vaccination coverage for the second dose of the measles-rubella vaccine remains low in this rural HD of Cameroon. The absence of an advanced strategy offers emerged as the main barrier to vaccination, regardless of other sociodemographic factors. These results call for a strengthening of the implementation of advanced strategies, which should be considered an essential lever for health equity in rural areas.

## Data Availability

All data produced in the present work are contained in the manuscript

## Abbreviations

*aOR*: Adjusted Odds Ratio
*cOR*: Crude Odds Ratio
*HD*: Health District
*MCV2*: Second Dose of the Measle Containing Vaccine

## Declarations

### Author contributions

Study design & conception: GSN and FZLC; Data collection: GSN and FZLC; Data analysis, visualization and interpretation: FZLC; Drafting of original manuscript: FZLC and GSN; Critical revision of the manuscript: FZLC, FKN, RMMA, and AA; Final approval of the manuscript: All authors.

### Ethical Approval Statement

Ethical clearance for the present study was waived by the Regional Delegation of Public Health Ethical Board. Administrative approval was obtained from the Chief Medical Officer of Ngo-Mapubi HD. Additionally, participants were required to provide signed informed consent. The confidentiality, anonymity and autonomy of the research participants were respected throughout the study. All methods were performed in accordance with the relevant guidelines of the Helsinki Declaration.

### Consent for publication

Not applicable.

### Availability of data and materials

All data generated or analyzed during this study are provided in the manuscript.

## Acknowledgments

Our gratitude goes to the Ngog-Mapubi community members for their hospitality during the conduction of this study.

