## Additional Files for "Community adherence to the second dose of measle and rubella: a cross-sectional survey in a rural health district of Cameroon"

**Supplementary Table 1** Reported MRV2 vaccination coverage among health areas of the Ngog-Mapubi HD, Cameroon (n = 140)

| **Health area*^1^*** | **MRV2 vaccination uptake**  **n (%)** | |
| --- | --- | --- |
|  | No  n = 75 (54) | Yes  n = 65 (46) |
| Bot-makak | 10 (90.9) | 1 (9.1) |
| Boumnyebel | 18 (60.0) | 12 (40.0) |
| Dibang | 4 (40.0) | 6 (60.0) |
| HEGBA | 1 (10.0) | 9 (90.0) |
| Mandoumba | 6 (54.5) | 5 (45.5) |
| Matomb | 3 (60.0) | 2 (40.0) |
| Mbanda | 3 (60.0) | 2 (40.0) |
| Mbebe-kikot | 3 (100.0) | 0 (0.0) |
| Mintaba | 3 (60.0) | 2 (40.0) |
| Ndongo | 5 (41.7) | 7 (58.3) |
| Ngog-Mapubi | 5 (35.7) | 9 (64.3) |
| Nguibassal | 2 (22.2) | 7 (77.8) |
| Ntouleng | 10 (90.9) | 1 (9.1) |
| Sombo | 2 (50.0) | 2 (50.0) |
| *^1^* Fisher exact probability test (*p*-value = 0.005); MRV2: second dose of the measles and rubella vaccine | | |

**Supplementary Table 2** Reported MRV2 vaccination acceptance among health areas of the Ngog-Mapubi HD, Cameroon (n = 75)

| **Health area*^1^*** | **MRV2 vaccination acceptance**  **n (%)** | |
| --- | --- | --- |
|  | No  n = 11 (15) | Yes  n = 64 (85) |
| Bot-makak | 0 (0.0) | 10 (100.0) |
| Boumnyebel | 3 (16.7) | 15 (83.3) |
| Dibang | 1 (25.0) | 3 (75.0) |
| HEGBA | 1 (100.0) | 0 (0.0) |
| Mandoumba | 0 (0.0) | 6 (100.0) |
| Matomb | 0 (0.0) | 3 (100.0) |
| Mbanda | 0 (0.0) | 3 (100.0) |
| Mbebe-kikot | 0 (0.0) | 3 (100.0) |
| Mintaba | 0 (0.0) | 3 (100.0) |
| Ndongo | 0 (0.0) | 5 (100.0) |
| Ngog-Mapubi | 2 (40.0) | 3 (60.0) |
| Nguibassal | 2 (100.0) | 0 (0.0) |
| Ntouleng | 2 (20.0) | 8 (80.0) |
| Sombo | 0 (0.0) | 2 (100.0) |
| *^1^* Fisher exact probability test (*p*-value = 0.047); MRV2: second dose of the measles and rubella vaccine | | |

**Supplementary Table 3** Reported compliance with the routine immunization program among health areas of the Ngog-Mapubi HD, Cameroon (n = 140)

| **Health area*^1^*** | **Adherence to EPI program**  **n (%)** | |
| --- | --- | --- |
|  | No  n = 9 (6) | Yes  n = 131 (94) |
| Bot-makak | 1 (9.1) | 10 (90.9) |
| Boumnyebel | 2 (6.7) | 28 (93.3) |
| Dibang | 0 (0.0) | 10 (100.0) |
| HEGBA | 0 (0.0) | 10 (100.0) |
| Mandoumba | 0 (0.0) | 11 (100.0) |
| Matomb | 0 (0.0) | 5 (100.0) |
| Mbanda | 0 (0.0) | 5 (100.0) |
| Mbebe-kikot | 0 (0.0) | 3 (100.0) |
| Mintaba | 0 (0.0) | 5 (100.0) |
| Ndongo | 3 (25.0) | 9 (75.0) |
| Ngog-Mapubi | 1 (7.1) | 13 (92.9) |
| Nguibassal | 2 (22.2) | 7 (77.8) |
| Ntouleng | 0 (0.0) | 11 (100.0) |
| Sombo | 0 (0.0) | 4 (100.0) |
| *^1^* Fisher exact probability test (*p*-value = 0.481); MRV2: second dose of the measles and rubella vaccine | | |

**Supplementary Table 4** Reported exposition to MRV2 communication activities among health areas of the Ngog-Mapubi HD, Cameroon (n = 140)

| **Health area*^1^*** | **Ever heard about MRV 2**  **n (%)** | |
| --- | --- | --- |
|  | No n = 15 (11) | Yes n = 125 (89) |
| Bot-makak | 4 (36.4) | 7 (63.6) |
| Boumnyebel | 3 (10.0) | 27 (90.0) |
| Dibang | 0 (0.0) | 10 (100.0) |
| HEGBA | 1 (10.0) | 9 (90.0) |
| Mandoumba | 1 (9.1) | 10 (90.9) |
| Matomb | 1 (20.0) | 4 (80.0) |
| Mbanda | 0 (0.0) | 5 (100.0) |
| Mbebe-kikot | 2 (66.7) | 1 (33.3) |
| Mintaba | 0 (0.0) | 5 (100.0) |
| Ndongo | 0 (0.0) | 12 (100.0) |
| Ngog-Mapubi | 1 (7.1) | 13 (92.9) |
| Nguibassal | 0 (0.0) | 9 (100.0) |
| Ntouleng | 1 (9.1) | 10 (90.9) |
| Sombo | 1 (25.0) | 3 (75.0) |
| *^1^* Fisher exact probability test (*p*-value = 0.062); MRV2: second dose of the measles and rubella vaccine | | |

**Supplementary Table 5** Reported exposition to MRV2 communication activities according to sociodemographic characteristics Ngog-Mapubi HD, Cameroon (n = 140)

| **Characteristic** | **Ever heard about MRV2**  **n (%)** | | ***p*-value*^1^*** |
| --- | --- | --- | --- |
|  | No n = 15 (11) | Yes n = 125 (89) |  |
| **Age group (years)** |  |  | >0.999 |
| ˂20 | 0 (0.0) | 7 (100.0) |  |
| 20-29 | 7 (11.9) | 52 (88.1) |  |
| 30-39 | 6 (10.7) | 50 (89.3) |  |
| 40+ | 2 (11.1) | 16 (88.9) |  |
| **Sex** |  |  | >0.999 |
| Female | 13 (10.9) | 106 (89.1) |  |
| Male | 2 (9.5) | 19 (90.5) |  |
| **Study level** |  |  |  |
| None | 4 (25.0) | 12 (75.0) |  |
| Primary | 5 (15.6) | 27 (84.4) |  |
| Secondary | 6 (6.9) | 81 (93.1) |  |
| Tertiary | 0 (0.0) | 5 (100.0) |  |
| **Marital status** |  |  | 0.360 |
| Single | 7 (12.3) | 50 (87.7) |  |
| Cohabitation | 5 (8.6) | 53 (91.4) |  |
| Married | 2 (8.7) | 21 (91.3) |  |
| Widow(er) | 1 (50.0) | 1 (50.0) |  |
| **Household’s member surveyed** |  |  | 0.118 |
| Grand-mother | 1 (100.0) | 0 (0.0) |  |
| Mother | 12 (10.2) | 106 (89.8) |  |
| Father | 2 (9.5) | 19 (90.5) |  |
| **Distance to the nearby health facility (km)** |  |  | 0.104 |
| 0-5 | 6 (7.1) | 78 (92.9) |  |
| 6+ | 9 (16.1) | 47 (83.9) |  |
| **Ethnic group** |  |  | 0.062 |
| Other tribes | 5 (22.7) | 17 (77.3) |  |
| Local tribe | 10 (8.5) | 108 (91.5) |  |
| **Occupation** |  |  | 0.879 |
| Civil servant | 0 (0.0) | 6 (100.0) |  |
| Farmer | 2 (8.3) | 22 (91.7) |  |
| Housekeeper | 8 (10.4) | 69 (89.6) |  |
| Other | 1 (11.1) | 8 (88.9) |  |
| Shopkeeper | 2 (16.7) | 10 (83.3) |  |
| Student | 2 (16.7) | 10 (83.3) |  |
| **Average monthly income (XAF)** |  |  | 0.725 |
| ˂50000 | 14 (11.6) | 107 (88.4) |  |
| 50000-150000 | 1 (5.6) | 17 (94.4) |  |
| ˃150000 | 0 (0.0) | 1 (100.0) |  |
| *^1^* Fisher exact probability test; MRV2: second dose of the measles and rubella vaccine; XAF: central African francs | | | |

| Reasons | Frequency (%) |
| --- | --- |
| Distance from vaccination site | 42.9 |
| Fever after vaccination | 14.3 |
| Lack of time | 14.3 |
| Lack of means of transport | 14.3 |
| Refusal of one of the legal guardian | 14.3 |

**Supplementary Fig. 1** Reported reasons for not usually getting vaccinated according to community members in the Ngog-Mapubi HD, Cameroon (n=9)

**Supplementary Table 6** Knowledge profile of community member regarding the MRV2 in the Ngog-Mapubi HD, Cameroon (n = 140)

| **Characteristic** | **Good knowledge**  **n (%)** | | ***p*-value*^1^*** |
| --- | --- | --- | --- |
|  | No n = 114 (81) | Yes n = 26 (19) |  |
| **Health area** |  |  |  |
| Bot makak | 10 (90.9) | 1 (9.1) |  |
| Boumnyebel | 28 (93.3) | 2 (6.7) |  |
| Dibang | 6 (60.0) | 4 (40.0) |  |
| HEGBA | 3 (30.0) | 7 (70.0) |  |
| Mandoumba | 10 (90.9) | 1 (9.1) |  |
| Matomb | 5 (100.0) | 0 (0.0) |  |
| Mbanda | 0 (0.0) | 5 (100.0) |  |
| Mbebe-kikot | 3 (100.0) | 0 (0.0) |  |
| Mintaba | 5 (100.0) | 0 (0.0) |  |
| Ndongo | 12 (100.0) | 0 (0.0) |  |
| Ngog Mapubi | 12 (85.7) | 2 (14.3) |  |
| Nguibassal | 9 (100.0) | 0 (0.0) |  |
| Ntouleng | 9 (81.8) | 2 (18.2) |  |
| Sombo | 2 (50.0) | 2 (50.0) |  |
| **Distance to the nearby health facility (km)** |  |  | 0.659 |
| 0-5 | 67 (79.8) | 17 (20.2) |  |
| 6+ | 47 (83.9) | 9 (16.1) |  |
| **Age group (years)** |  |  | 0.827 |
| ˂20 | 6 (85.7) | 1 (14.3) |  |
| 20-29 | 50 (84.7) | 9 (15.3) |  |
| 30-39 | 44 (78.6) | 12 (21.4) |  |
| 40+ | 14 (77.8) | 4 (22.2) |  |
| **Sex** |  |  | 0.544 |
| Female | 98 (82.4) | 21 (17.6) |  |
| Male | 16 (76.2) | 5 (23.8) |  |
| **Marital status** |  |  | 0.896 |
| Single | 46 (80.7) | 11 (19.3) |  |
| Cohabitation | 48 (82.8) | 10 (17.2) |  |
| Married | 18 (78.3) | 5 (21.7) |  |
| Widow(er) | 2 (100.0) | 0 (0.0) |  |
| **Study level** |  |  | 0.097 |
| None | 15 (93.8) | 1 (6.3) |  |
| Primary | 29 (90.6) | 3 (9.4) |  |
| Secondary | 65 (74.7) | 22 (25.3) |  |
| Tertiary | 5 (100.0) | 0 (0.0) |  |
| **Occupation** |  |  | 0.694 |
| Civil servant | 5 (83.3) | 1 (16.7) |  |
| Farmer | 19 (79.2) | 5 (20.8) |  |
| Housekeeper | 64 (83.1) | 13 (16.9) |  |
| Other | 7 (77.8) | 2 (22.2) |  |
| Shopkeeper | 8 (66.7) | 4 (33.3) |  |
| Student | 11 (91.7) | 1 (8.3) |  |
| **Household’s member surveyed** |  |  | 0.372 |
| Grand-mother | 1 (100.0) | 0 (0.0) |  |
| Mother | 98 (83.1) | 20 (16.9) |  |
| Father | 15 (71.4) | 6 (28.6) |  |
| **Household’s number of children** |  |  | 0.762 |
| 0-3 | 43 (84.3) | 8 (15.7) |  |
| 3-5 | 43 (81.1) | 10 (18.9) |  |
| 5-8 | 28 (77.8) | 8 (22.2) |  |
| **Household’s number of 15-23 months old children** |  |  | >0.999 |
| 0-1 | 102 (81.0) | 24 (19.0) |  |
| 2+ | 12 (85.7) | 2 (14.3) |  |
| *^1^* Fisher exact probability test | | | |
